## Supplementary Material for "Empowering Refugee Voices: Using Nominal Group Technique with a Diverse Refugee Patient Advisory Committee (PAC) to Identify Health and Research Priorities"

**Supplementary Materials:**

eTable 1: Dates of focus group meetings and their topic of discussion.

eTable 2a. Raw rankings of research priorities for pre-migration/early arrival time-period.

eTable 2b. Grouped-together priorities with similar ideas and their combined total votes.

eTable 2c. Summarized similar priorities into a single sentence with their combined votes.

eTable 2d. Final concise one sentence summary priorities for the research priorities for pre-migration/early arrival time-period (0 – 3 months).

**APPENDIX**

eTable 1. Dates of focus group meetings and their topic of discussion

| Data of PAC Session | Discussion Topic(s) |
| --- | --- |
| January 17^th^, 2023 | **research priorities** for **post-migration period** (up to 2 years). - issues of most importance for refugee health research |
| March 14^th^, 2023 | **health priorities** for **pre-migration/early arrival** period and **post-migration** (up to 2 years) period. |
| May 16^th^, 2023 – Ukrainian Only | **health and research** priorities for the **post-migration** (up to 2 years) period. |
| November 16^th^, 2023 | **health and research** priorities for l**ong term resilient** health system. |
| January 29^th^, 2024 | **research priorities** for the **pre-migration/early arrival** period and prioritized **top 5 across all** health and research priorities. |

eTable 2a. Raw rankings of research priorities for pre-migration/early arrival time-period.

| **Priorities** | Total |
| --- | --- |
| What are the most important things that refugees must be aware of when it comes to Canadian healthcare system prior to arrival | **12** |
| How can I navigate/leverage w the education system (ie. Linc classes) to promote healthcare system info, navigation | **12** |
| How can I find out info about medical expenses prior to arrival so I can plan pre-departure? | **12** |
| How can we help make healthcare information more available for refugees post first arrival? | **10** |
| What was your greatest need for your health when you first arrived? | **9** |
| How do I navigate health system? And who can teach me? | **9** |
| Are ppl eligible to receive medication that is important or critical that they use at home in Canada? | **8** |
| Health system orientation pre-departure is critical how | **8** |
| Can Canada introduce health system navigation awareness classes (ie., like mandatory language instruction classes) for health so that all new refugees receive? | **7** |
| How can gov provide a short and complete information that is understandable for post arrival refugees re: vaccinations (routine vs. other) and why important | **7** |
| How do we improve access to pre-departure medical records from home country to Canadian providers | **7** |
| What did you worry about the most before you arrived in Canada? | 5 |
| Upon arrival – how do refugees obtain info about clinics that they can go to? | 5 |
| What to expect when you arrive re: housing, job search, HC, schools, social services and education? | 5 |
| How to deal w pre-existing conditions post arrival? | 5 |
| Possible ways to provide feedback for the IME exams? (i.e., did the clinics submit this information to IRCC?) | 5 |
| Do children or youth receive any assessments re: MH post arrival? Do they receive any access to care, counselling, treatments? | 5 |
| Important health info pre-arrival that is critical for post arrival providers to be safe? How do we make this info available (ie. Woman that is 9months pregnant at departure, is unknown to HC providers post arrival) | 5 |
| IFHP coverage and provincial HC card. How do we improve understanding of both programs and what covers what? | 5 |
| Families are unprepared for the challenges that their children are facing or will face in Canada. How do we support new arrival parents to help and/or support their children (1st gen kids facing issues not common in COO)? | 5 |
| What are the most common or most important health conditions for refugees by global region? | 3 |
| How does finding a job or employment status affect the mental health of newcomers post arrival? | 3 |
| What is the prevalence of communicable and NCDs pre-arrival and how does this differ by global region | 3 |
| How do we improve healthcare providers’ training in Canadian HC system to increase capacity in general | 3 |
| What is the prevalence of MH d/o for children camps vs. no-camps and COO and Region OO? | 2 |
| Question How much to you know about the Canadian HC system? (Question to refugees prior to departure) | 2 |
| How can I communicate w the HC system if I see a violation of conduct by HC employees or institutions? | 2 |
| How can I get health assessment and make me understand it (or results)? | 0 |
| How to IME exam information requirements more clear for refugees pre/post arrival? | 0 |
| Many ppl may be considering coming to Canada as refugees but have health conditions that require HC. How to make info clear whether pre-existing Health conditions would exclude one from coming to Canada? | 0 |
| Why should I receive these vaccinations after arrival? | 0 |

eTable 2b. Grouped-together priorities with similar ideas and their combined total votes.

| **Priorities after combination of similar ideas** | Votes | Combined votes |
| --- | --- | --- |
| How can I navigate/leverage w the education system (ie. Linc classes) to promote healthcare system info, navigation | 12 | 55 |
| How can we help make healthcare information more available for refugees post first arrival? | 10 |  |
| How do I navigate health system? And who can teach me? | 9 |  |
| Can Canada introduce health system navigation awareness classes (ie., like mandatory language instruction classes) for health so that all new refugees receive? | 7 |  |
| Upon arrival – how do refugees obtain info about clinics that they can go to? | 5 |  |
| IFHP coverage and provincial HC card. How do we improve understanding of both programs and what covers what? | 5 |  |
| How can I get health assessment and make me understand it (or results)? | 0 |  |
| How to IME exam information requirements more clear for refugees pre/post arrival? | 0 |  |
| Why should I receive these vaccinations after arrival? | 0 |  |
| How can gov provide a short and complete information that is understandable for post arrival refugees re: vaccinations (routine vs. other) and why important | 7 |  |
| What are the most important things that refugees must be aware of when it comes to Canadian healthcare system prior to arrival | 12 | 42 |
| Health system orientation pre-departure is critical how | 8 |  |
| How much to you know about the Canadian HC system? (Question to refugees prior to departure) | 2 |  |
| How can I find out info about medical expenses prior to arrival so I can plan pre-departure? | 12 |  |
| Are ppl eligible to receive medication that is important or critical that they use at home in Canada? | 8 |  |
| Many ppl may be considering coming to Canada as refugees but have health conditions that require HC. How to make info clear whether pre-existing Health conditions would exclude one from coming to Canada? | 0 |  |
| Important health info pre-arrival that is critical for post arrival providers to be safe? How do we make this info available (ie. Woman that is 9months pregnant at departure, is unknown to HC providers post arrival) | 5 | 17 |
| How to deal w pre-existing conditions post arrival? | 5 |  |
| How do we improve access to pre-departure medical records from home country to Canadian providers | 7 |  |
| What was your greatest health needs for your health when you first arrived? | 9 | 14 |
| What did you worry about the most before you arrived in Canada with regards to health? | 5 |  |
| Do children or youth receive any assessments re: MH post arrival? Do they receive any access to care, counselling, treatments? | 5 | 12 |
| What is the prevalence of MH d/o for children camps vs. no-camps and COO and Region OO? | 2 |  |
| Families are unprepared for the challenges that their children are facing or will face in Canada. How do we support new arrival parents to help and/or support their children (1st gen kids facing issues not common in COO)? | 5 |  |
| Possible ways to provide feedback for the IME exams? (i.e., did the clinics submit this information to IRCC?) | 5 | 7 |
| How can I communicate w the HC system if I see a violation of conduct by HC employees or institutions? | 2 |  |
| What are the most common or most important health conditions for refugees by global region? | 3 | 6 |
| What is the prevalence of communicable and NCDs pre-arrival and how does this differ by global region | 3 |  |
| What to expect when you arrive re: housing, job search, HC, schools, social services and education? | 5 | 5 |
| How does finding a job or employment status affect the mental health of newcomers post arrival? | 3 | 3 |
| How do we improve healthcare providers’ training in Canadian HC system to increase capacity in general | 3 | 3 |

eTable 2c. Summarized similar priorities into a single sentence with their combined votes.

| One sentence summary | Votes |
| --- | --- |
| 1. How can we make health system information more available or leverage existing programs (i.e. language Classes) to increase health system navigation **post arrival**, including: clinics to go to, vaccinations, insurance coverage, resources, healthcare processes, immigration medical exam, where to start and the importance of each | 55 |
| 2. How can we enhance health system navigation **pre-arrival** as it is critical, which includes: medical expenses orientation, health conditions transition treatments (and whether or not health conditions can affect migration status negatively), and crucial orientation points that refugees should know prior to migration. | 42 |
| 3. How can Canada improve health data transfer in terms of access to medical records from home country and health status and needs to Canadian health system immediately post arrival (ie. Woman that is 9 months pregnant at departure, is unknown to HC providers post arrival). | 17 |
| 4. what were your greatest needs and worries in regards to health prior to coming to Canada? | 14 |
| 5. What is the prevalence of mental health issues for children in camps vs. no-camps and COO and Region OO, and do children or youth receive mental health assessments, care and treatment post arrival and how are parents supported in this regard? | 12 |

eTable 2d. Final concise one sentence summary priorities for the research priorities for pre-migration/early arrival time-period (0 – 3 months).

| One sentence summary | Votes |
| --- | --- |
| How to create/improve post-arrival refugee healthcare system navigation? | 55 |
| How to improve pre-arrival refugee healthcare guidance/navigation? | 42 |
| How to ensure pre --> post migration refugee health info continuity sharing? | 17 |
| What are refugees’ key pre-arrival health concerns/worries/needs/expectations? | 14 |
| How to provide refugee children’s mental health & parental supports? | 12 |
